# Success factors of growth-stage digital health companies: protocol for a systematic literature review

**DOI:** 10.1101/2024.05.06.24306674

**Authors:** Estelle Pfitzer, Laura Bitomsky, Marcia Nißen, Christoph Kausch, Tobias Kowatsch

## Abstract

**Background:** The healthcare industry is undergoing rapid transformation and progress, fueled by technological advances and the growing need for accessible, affordable, and high-quality healthcare services. While there has been significant progress in digitizing healthcare, especially during the COVID-19 pandemic, there is still considerable untapped potential for digital therapeutics (DTx). According to a report by Bloomberg, the digital health market is projected to reach $1.5 trillion by 2030 (1). However, most ventures in the space struggle to gain traction and eventually stop operations. Also, the diverse regulatory guidelines across countries challenge the scalability of DTx.

**Objective:** The protocol described here outlines the methodology for a systematic review which’s objective is to identify factors that contribute to the successful scaling of digital health companies that have a validated product or service, i.e., growth-stage companies. A further objective is to highlight the key metrics used to quantify success in digital health companies.

**Method:** We will search business and scientific databases, including EBSCO, PubMed, ProQuest, and Scopus, summarize the key findings and highlight the specific success factors relevant to the Digital Health Technologies (DHT) industry. The quality of the selected studies will be assessed using the critical appraisal skills program (CASP) checklists.

**Expected outcome:** The literature review’s primary objective is to identify essential characteristics and recurring patterns that significantly contribute to success among growth-stage digital health companies. It will bridge the existing knowledge gap and provide stakeholders, including investors and entrepreneurs, with a valuable resource that supports informed decision-making and enhances the success of growth-stage digital health companies.

**OSF Registration:** https://doi.org/10.17605/OSF.IO/UYJCA

## Introduction

The past decade saw a remarkable and rapid advancement in the development of digital health technologies. The COVID-19 pandemic propelled the adoption of these technologies, leading to significant innovation and supportive regulatory reforms. In the same period, investor interest surged, and investments in digital health companies grew more than tenfold (2,3) accelerating solutions that promise to disrupt and reshape the landscape of modern medicine, making care more accessible, efficient, and affordable (4,5).

In the dynamic landscape of digital health, however, many companies fail to sustain momentum beyond early-stage successes, even when initially considered unicorns. For example, Pear Therapeutics, the first company to receive FDA approval for its standalone digital therapeutics (DTx) solution, reSET®, in September 2017. reSET® is a prescription DTx designed to treat substance use disorder in conjunction with outpatient counseling (6). Pear Therapeutics secured substantial funding, reaching a valuation of more than $1.5 billion (7), and earning a position on several lists of leading DTx companies (8). And yet, in 2023, the company ultimately filed for bankruptcy, providing a cautionary tale for the entire sector.

Proteus Digital Health also made headlines in 2020 when it filed for bankruptcy. The company had reached a significant milestone by developing the first FDA-approved “smart pill” capable of monitoring medication intake and tracking its effectiveness. Its innovative approach has been rewarded with a valuation of $1.5 billion. But despite the technology’s validation through studies and clinical trials, the company faced challenges in scaling and gaining substantial market traction (9).

In October 2020, Aidhere launched Zanadio, the third digital health application (DiGA), to receive approval from the German Federal Institute for Drugs and Medical Devices (BfArM). Zanadio made history as the first digital obesity therapy on prescription to be eligible for insurance reimbursement in Germany. Despite having the top-performing (10) digital health product on the DiGA directory and successfully treating over 30’000 overweight patients, Aidhere filed for bankruptcy in May 2023. The company struggled to reach an agreement with insurance companies, which resulted in a drastic reduction in the price of their therapy, rendering the business model unsustainable. This struggle is not unique to Aidhere, as almost all other 52 DiGAs are confronted with similar obstacles and are now compelled to seek alternative solutions.

Even the most revolutionary DTx cannot significantly impact patients’ lives if they fail to align with the interests of healthcare payers, such as insurers, national healthcare systems, and employers who offer health benefits to their employees.

Corey McCann, a former CEO of Pear Therapeutics, stated on the day of announcing insolvency:

“We’ve shown that clinicians will readily prescribe PDTs [prescription DTx, the authors]. We’ve shown that patients will engage with the products. We’ve shown that our products can improve clinical outcomes. We’ve shown that our products can save payors money. Most importantly, we’ve shown that our products can truly help patients and their clinicians. But that isn’t enough. Payors have the ability to deny payment for therapies that are clinically necessary, effective, and cost saving. In addition, market conditions over the last two years have challenged many growth-stage companies, including us.”

According to the Global Strategy on Digital Health by the World Health Organization (11), the success of digital health technologies (DHTs) relies on ensuring accessibility: DHTs should improve the efficiency and sustainability of health systems, delivering care that is both affordable and equitable. They should also play a role in strengthening and expanding health promotion, disease prevention, diagnosis, management, rehabilitation, and palliative care. And DHTs need to prioritize the privacy and security of patients’ health information. While previous studies have examined success factors within various industries (12,13), there is a notable gap in research focusing specifically on digital health companies. This review aims to bridge that gap by comprehensively analyzing general success factors applicable to growth-stage digital companies and digital health-specific success factors. We will further focus specifically on the different categories within the DHT industry as defined by the DTx Alliance (14). These categories include patient-facing solutions such as Digital Therapeutics, Digital Diagnostics, Care Support, Patient Monitoring, and Wellness, as well as software solutions for healthcare providers and other digital health stakeholders like pharmaceutical companies.

To this end, we formulate the following research questions (RQs):

RQ1: What are the general success factors of growth-stage digital companies?

RQ2: What are the specific success factors of growth-stage digital health companies?

RQ3: How do digital health companies’ success factors differ between the DHT categories?

To address these research questions and based on prior research (12,13,15–20), we will identify factors that are directly related to the companies’ success.

## Methods

We conduct a systematic literature review to answer our RQs, and thus, assess systematically the fragmented knowledge base on success factors in growth-stage digital (health) companies. This review follows established guidelines proposed by Snyder and Tranfield et al. (21,22), ensuring the generation of evidence-informed management knowledge. To this end, appropriate elements from the Preferred Reporting Items for Systematic reviews and Meta-Analyses (PRISMA) statement are considered (23).

### Eligibility criteria

We have meticulously established inclusion and exclusion criteria for the systematic literature review, which focuses on examining the success factors of growth-stage digital (health) companies. The inclusion and exclusion criteria are outlined in Tables 1 and 2, respectively. To ensure a comprehensive analysis of the subject matter, we have included papers published from 2000 to 2023. This time frame was selected as the early 2000s marked a significant period in the history of digital companies, known as the burst of the Dot-Com Bubble. This era was characterized by a notable influx of internet-based or digital companies entering the market (24).

**Table 1:** Inclusion Criteria.

| Inclusion Criteria | Reason for inclusion |
| --- | --- |
| Research focus | Studies that identify the success factors in digital growth-stage companies |
| Year | Only studies from 2000 and on are considered |
| Language | Only English studies are considered |
| Publication type | Peer reviewed academic literature |

**Table 2:**
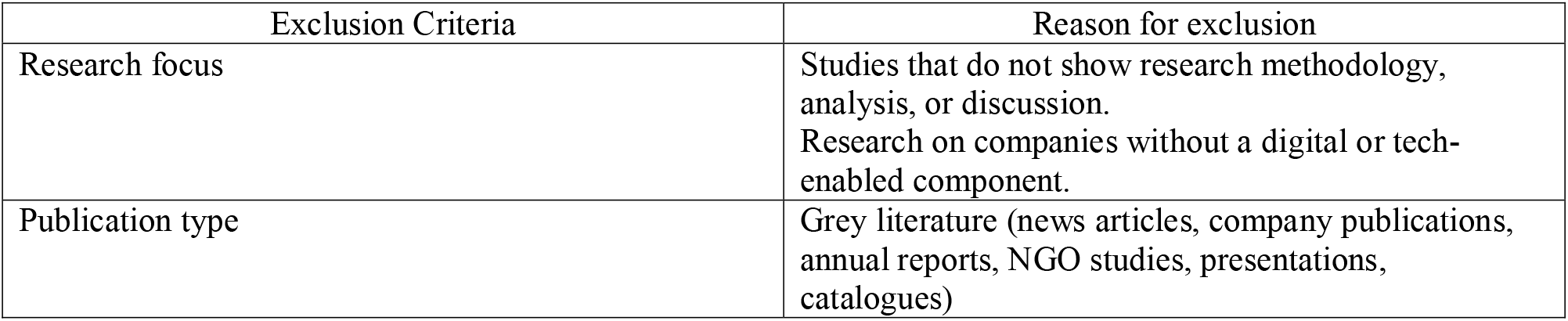
Exclusion Criteria.

| Exclusion Criteria | Reason for exclusion |
| --- | --- |
| Research focus | Studies that do not show research methodology, analysis, or discussion.<br>Research on companies without a digital or tech-enabled component. |
| Publication type | Grey literature (news articles, company publications, annual reports, NGO studies, presentations, catalogues) |

### Information Sources

To ensure a comprehensive search, we will use the following databases:

- EBSCO and ProQuest: These databases are widely recognized and commonly used for business research, offering a diverse range of business-related journals, articles, and reports.
- PubMed: This database specializes in biomedical and life sciences research, providing valuable insights at the intersection of healthcare and business.
- Scopus and Web of Science: As multidisciplinary databases, they cover a broad spectrum of subject areas, enabling us to obtain comprehensive coverage of our topic.

By exploring these databases, we will gather diverse sources covering business-specific, healthcare-business intersections, and interdisciplinary perspectives, ensuring a thorough search for our study.

### Search Strategy

The systematic review utilized specific search terms listed in Table 3. These terms are employed to search various databases, including the publications’ titles, abstracts, and keyword sections. The search terms were carefully chosen based on relevant words, concepts, and synonyms that are closely related to the research questions (22). To retrieve the relevant literature, we will conduct the search in the databases using the search terms I AND II AND III AND IV. Additionally, for RQ2 and RQ3 specific to digital health, search terms V will be specifically searched within the full text of selected publications. For the exact search strings used in each database, please refer to the Appendix. This approach ensures a systematic and comprehensive retrieval of relevant literature for the review.

**Table 3:** Search terms based on alternative keywords.

| NB | Keyword category | Search terms |
| --- | --- | --- |
| I | Success Factors | Success factors* OR Key success factors* OR Critical success factors* OR Factors contributing to success* OR Determinants of success* OR Drivers of success* OR Elements of success* OR Factors influencing success* OR Achieving success* OR Factors for successful outcomes* OR Achieving desired outcomes* OR Success determinants* OR Successful outcome* OR Predicted outcome* OR Outcomes of success* OR Desired outcome* OR Outcome measurement* OR Outcome evaluation* OR Outcome assessment* OR High performance* OR Performance* OR Success prediction* |
| II | Growth-stage | Growth stage* OR Development stage* OR Expansion stage* OR Scale-up stage* OR Scale-up* OR Scaling* OR High-growth stage* OR Rapid growth stage* OR Growth-oriented* OR Growing companies* OR Maturing companies* OR Evolving companies* OR Advancing companies* OR Progressive companies* OR Progressing* |
| III | Companies | Companies* OR Firms* OR Organizations* OR Enterprises* OR Ventures* OR Startups* OR Businesses* OR Industry* |
| IV | Digital | Digital* OR Digitization* OR Digitalization* OR Digital transformation* OR Digital solutions* OR Digital technologies* OR Digital tools* OR Digital systems* OR Digital processes* OR Digital capabilities* OR Digital strategies* OR Digital adoption* OR Digital implementation* OR Technology-driven* OR Tech-enabled* OR Tech-oriented* OR Tech-driven* OR Technology-focused* OR Software* OR Software solution* OR Automation* OR E-commerce* OR Internet of Things* OR Cloud computing* OR Data analytics* OR Cybersecurity* OR Artificial Intelligence* OR AI* OR Machine Learning* OR ML* OR Mobile apps* OR Web applications* OR User experience* OR Platform* OR Connectivity* OR Digital innovation* OR Digital disruption* |
| V | Digital Health | Health* OR Medicine* OR Digital health* OR Health technology* OR |
|  |  | Medical technology* OR Health IT* OR Healthcare technology* OR Health informatics* OR Telemedicine* OR Telehealth* OR Health apps* OR Wearables* OR E-health* OR M-health* OR Health innovation* OR Healthtech* OR MedTech* OR Health data* OR Health analytics* OR Health software* OR Health platforms* OR Medical informatics* OR Healthcare informatics* OR Digital medicine* OR Remote patient monitoring* OR Virtual healthcare* OR Health monitoring technology* OR Mobile health application* OR Health data management* OR Wearable health device* OR Digital therapeutic* OR Health information system* OR Healthcare informatic* OR Remote health monitoring* OR Electronic health records* OR Connected health devices* OR Health tech advancements* OR Health technology integration* OR Health technology adoption* OR Health technology assessment* OR Health technology implementation* OR Healthcare software solution* OR Mobile medical application* |

### Study Records

#### Data management

After conducting comprehensive searches in relevant databases, the obtained citations will be imported into Mendeley 2.91.0, a reference management software. All extracted data and supporting documents will be securely stored in Mendeley to ensure data integrity and facilitate collaboration.

#### Selection process

The selection process will involve pairs of co-authors independently assessing the titles and abstracts with Rayyan (https://www.rayyan.ai/) to determine their eligibility based on the inclusion criteria. The lead author (EP) will obtain the full-text articles, which pairs of authors will review for potential inclusion to minimize bias in study selection. Any discrepancies during the screening process will be resolved through discussion between the authors or by seeking the opinion of a third co-author. The selection procedure will be meticulously documented, including creating a PRISMA flow diagram and a list of excluded studies with reasons for their exclusion. Additionally, a separate list of single-center studies initially excluded during the title and abstract screening but meeting the inclusion criteria will be recorded and available upon request.

Publications selected for RQ1, related to digital health will be used to answer RQ2 and RQ3. As described above, this secondary selection process will also be conducted systematically, involving two independent reviewers who will collaborate and address any discrepancies.

#### Data collection process

A standardized data extraction form will be developed and utilized to collect relevant information for this systematic literature review on the success factors of growth-stage digital health companies. The form will be piloted on a subset of studies (2-3) to ensure its effectiveness and refine any necessary modifications.

Pairs of authors will independently extract data from the included studies. In case of discrepancies, these will be resolved through discussions between the authors. If any disagreements persist, a third author will be consulted. This rigorous approach ensures the reliability and accuracy of the data extraction process.

#### Risk of bias and quality assessment

The Critical Appraisal Skills Programme (CASP) checklist for qualitative research will be used to assess the quality of the selected studies, with a focus on identifying and considering the impact of any methodological limitations on our research outcomes (25). CASP is not only a widely recognized tool but also endorsed by authoritative bodies like Cochrane and the World Health Organisation for its effectiveness in qualitative evidence synthesis (25–27). This comprehensive tool examines a study’s methodological strengths and weaknesses through ten targeted questions, covering aspects ranging from clarity of aim to the overall significance and contribution of the study.

To ensure unbiased and thorough appraisal, two independent authors will evaluate each study. They will use the CASP framework to assign ratings of “Yes”, “Can’t tell”, or “No” to each question. A “No” response to any methodological question will indicate poor quality. In case of disagreements, a discussion between reviewers will be initiated to reach consensus. If needed, a third reviewer will be consulted to provide additional perspective and help resolving any persisting discrepancies.

#### Data Items

We will extract and analyze specific strategies, collaborative efforts, and organizational practices from various digital companies that have been demonstrated to positively influence business performance, encompassing areas such as external environments and relationships, business strategies and market positioning, as well as operational structures. We will categorize the success variable into groups to facilitate comparisons across different studies. The selection of these early groups is based on relevant papers and reports from early exploratory research (8,11,12,20,28). We have also provided examples of variables that fit into each category. These segments will be validated through our review process, and additional groups will be included to cover all researched success factors. This segmentation approach will enable a deeper understanding of which aspects and strategic focuses of companies are most crucial in driving the scaling and growth of digital and specifically digital health companies. A comprehensive table will be included to summarize the identified factors in each study, providing a clear overview of the most critical components contributing to the successful growth of digital and digital health firms.

Team Composition:

- Experience in the field (measured in years)
- Previous founding experience
- Academic formation of the team (qualifications and degrees)
- Gender distribution
- Age of the founding team
- Diversity of skills and expertise within the team
- Leadership experience and qualities of team members

Product/Service:

- Number of clinical trials conducted by the company
- Number of patents held by the company
- Product innovation
- Market demand and relevance of the product/service
- Quality and performance of the product/service
- Intellectual property protection
- Customer feedback and satisfaction with the product/service

Effectiveness of Strategic Partnerships:

- Early government support
- Amount of grants received
- Board composition
- Investors related factors
- Synergy and alignment between partners’ goals and objectives
- Collaborative innovation and knowledge sharing with partners
- Access to resources, networks, and expertise through partnerships

Geographic Locations:

- Regulatory environment of the geographic location
- Dynamism of the environment (presence of a vibrant and supportive ecosystem)
- Presence in strategic healthcare markets or regions
- Access to local talent pools and specialized resources (distance to top universities)
- Supportive regulatory framework for digital health companies
- Proximity to potential customers, investors, or partners

Business Models:

- Revenue generation strategies and models (e.g., subscription-based, transactional, or advertising-based)
- Value proposition and differentiation in the market
- Adaptability and flexibility to market changes and disruptions

Competitive Landscape:

- Number of competitors in the market
- Competitive advantage or unique selling proposition
- Market share and customer acquisition rates compared to competitors
- Innovation and differentiation in products/services compared to competitors
- Collaborative opportunities with competitors for mutual growth

Impact on health care efficiency:

- Reduction of administrative burden
- Improved accuracy and speed of diagnosis
- Enhanced coordination and communication among providers
- Resource allocation and utilization optimization

Impact on health care accessibility:

- Increase in Affordability
- Expanded reach to underserved areas
- Improved healthcare infrastructure and facilities
- Integration with existing systems
- Reduction of barriers to accessing care

Stakeholders’ involvement:

- Patient and healthcare professional engagement
- Support from regulatory bodies and policymakers
- Collaboration with insurance providers and healthcare organizations
- Incorporation of patient feedback and preferences

### Outcomes

#### Main outcome

Our research aims to generate a comprehensive list of success factors for growth-stage digital companies, ranked by the number of times they have been identified in selected studies. This will provide insights into the relative importance and prevalence of these success factors in the growth and scalability of digital companies. In addition, our focus on RQ2 and RQ3 specifically targets growth-stage digital health companies, allowing us to pinpoint factors that hold particular significance in the healthcare sector.

#### Additional outcomes

Comparative Analysis of Success Factors: Our investigation will compare the success factors specific to growth-stage digital health companies with those in non-health-related fields. This analysis will reveal the unique variables critical for success within the digital health environment. We will specifically categorize DHT companies to reveal the distinctive factors contributing to scaling in each category. Doing so will give us a deeper understanding of the diverse dynamics and requirements within the digital health landscape.

Evaluation of Success Metrics: As part of our comprehensive data collection process, we will gather information on various success metrics utilized in the digital health industry. These metrics include revenue growth, market share, user adoption, and investor funding. Through careful analysis, we will gain insights into the different indicators of success employed in the industry. This evaluation will highlight the key metrics used to measure and quantify success in digital companies.

### Data Synthesis

The findings of this systematic review will be presented in a narrative summary, accompanied by the Preferred Reporting Items for Systematic Reviews and Meta-analyses (PRISMA) 2020 flow diagram to depict the review process. To provide an overview of the included studies,, we will employ a detailed table. This table will systematically display key information for each study, including the study’s title, its specific sector within digital technologies, the definition of success employed, and the methodology used to identify success factors.

In addition, we will create a second table designed for cross-referencing purposes. This table will feature the papers on the y-axis and the categorized success factors on the x-axis. It will effectively highlight and compare the key characteristics and findings of each paper relative to the identified success factors. This tabular representation will facilitate an easy comparison and synthesis of data across the various studies, providing a clear visual summary of the overlapping and unique elements in each.

## Conclusion

The methodically curated list of success factors will serve as valuable resource for entrepreneurs, investors, and stakeholders operating in the digital industry, particularly within the digital health domain. This resource will empower them to prioritize and focus on the most influential factors for success in this dynamic and competitive market. Especially, founders in the digital health domain who have an established product or service and have acquired their first paying customers but struggle to transition from initial traction to significant scaling, will find this research invaluable in providing targeted guidelines on which specific strategies, collaborative efforts, and organizational practices to emphasize.

Investors in growth-stage digital health companies will find this research instrumental not only in making more informed decisions about which companies have set the right priorities and are more likely to scale successfully, but also in providing structured recommendations on effective scaling strategies for digital health businesses, particularly when serving on their boards. This dual benefit enhances both investment decision-making and strategic guidance capabilities.

Furthermore, our systematic review will not only synthesize existing knowledge on success factors in digital health startups but also shed light on the gaps and limitations present in the current literature and can guides future research directions. Ultimately, our findings establish a solid foundation for future studies, facilitating a more comprehensive understanding of the success factors that propel growth in the digital health industry.

## Data Availability

All data produced in the present work are contained in the manuscript

## Author’s contribution

All authors played integral roles in shaping this protocol and its revisions. TK and EP conceived the search questions, while EP formulated the review approach and design with inputs from TK and CK. EP initiated the manuscript drafting, incorporating valuable feedback from TK throughout the iterations. LB will support in the coding of the literature review. The final version received full approval from all authors.

## Supporting Information

S1 File. Prisma checklist

## References

1. Digital Health Market, Inc. - Bloomberg [cited 2023 Jul 13]. Available from: https://www.bloomberg.com/press-releases/2022-10-14/digital-health-market-to-hit-1-5-trillion-by-2030-grand-view-research-inc

2. 2021 digital health funding - Rock Health 2022. Available from: https://rockhealth.com/insights/2021-year-end-digital-health-funding-seismic-shifts-beneath-the-surface/

3. 2022 year-end digital health funding: Lessons at the end of a funding cycle Rock Health 2023. Available from: https://rockhealth.com/insights/2022-year-end-digital-health-funding-lessons-at-the-end-of-a-funding-cycle/

4. Meskó B, Drobni Z, Bényei É, Gergely B, Győrffy Z. Digital health is a cultural transformation of traditional healthcare. Mhealth 2017 Sep;3:38. doi: 10.21037/mhealth.2017.08.07

5. Mathews SC, McShea MJ, Hanley CL, Ravitz A, Labrique AB, Cohen AB. Digital health: a path to validation. npj Digital Medicine 2019 2:1 2019;2(1):1–9. doi: 10.1038/s41746-019-0111-3

6. FDA permits marketing of mobile medical application for substance use disorder | FDA [cited 2023 Jul 13]. Available from: https://www.fda.gov/news-events/press-announcements/fda-permits-marketing-mobile-medical-application-substance-use-disorder

7. SoftBank-backed Pear Therapeutics agrees $1.6 billion SPAC deal. Reuters 2021 Jun; Available from: https://www.reuters.com/article/us-pear-therapeutics-spac-idUSKCN2DY158

8. Hong JS, Wasden C, Han DH. Introduction of digital therapeutics. Comput Methods Programs Biomed 2021 Sep;209:106319. doi: 10.1016/j.cmpb.2021.106319

9. Litvinova O, Klager E, Tzvetkov NT, Kimberger O, Kletecka-Pulker M, Willschke H, et al. Digital Pills with Ingestible Sensors: Patent Landscape Analysis. Pharmaceuticals 2022, Vol 15, Page 1025 2022 Aug 19 [cited 2023 Jul 13];15(8):1025. doi: 10.3390/ph15081025

10. GKV-Spitzenverband. Bericht des GKV-Spitzenverbandes über die Inanspruchnahme und Entwicklung der Versorgung mit digitalen Gesundheitsanwendungen (DiGA-Bericht) gemäß § 33a Absatz 6 SGB V.

11. World Health Organization 2021. Global strategy on digital health 2020-2025 Geneva; [cited 2023 Jul 13]. Available from: https://www.who.int/docs/default-source/documents/gs4dhdaa2a9f352b0445bafbc79ca799dce4d.pdf

12. Santisteban J, Mauricio D. Systematic literature review of critical success factors of Information Technology startups. Academy of Entrepreneurship Journal. 2017 Nov;23:1–23.

13. Skawińska E, Zalewski RI. Success Factors of Startups in the EU—A Comparative Study. Sustainability 2020;12(19):8200. doi: 10.3390/su12198200

14. https://www.dtxalliance.org FACT SHEET Digital Health Technology Ecosystem Categorization What Are Digital Health Technologies? [cited 2023 Aug 11]; Available from: https://www.dtxalliance.org

15. Fitria SE, Hakim FR. Identification of Critical Success Factor Startup in Business Incubators (Case Study: Bandung Techno Park). International Journal of Social Service and Research 2022; 2(10):881–95. doi: 10.46799/ijssr.v2i10.162

16. Iyer AK. Building Digital Health and Therapeutic Solutions for the Future: What’s Required for Success? Digital Therapeutics: Strategic, Scientific, Developmental, and Regulatory Aspects 2022 Jan 1 [cited 2023 Jul 19];245–60. eBook ISBN: 9781003017288

17. Werth O, Cardona DR, Torno A, Breitner MH, Muntermann J. What determines FinTech success?—A taxonomy-based analysis of FinTech success factors. Electronic Markets 2023; 33(1). doi: 10.1007/s12525-023-00626-7

18. Skawinska E, Zalewski RI. Success Factors of Startups in the EU—A Comparative Study. Sustainability 2020, Vol 12, Page 8200 2020;12(19):8200. doi: 10.3390/su12198200

19. Santisteban J, Mauricio D, Cachay O. Critical success factors for technology-based startups. International Journal of Entrepreneurship and Small Business 2021;42(4):397–421. doi: 10.1504/IJESB.2021.114266

20. Zbikowski K, Antosiuk P. A machine learning, bias-free approach for predicting business success using Crunchbase data. Inf Process Manag. 2021 Jul 1;58(4):102555.

21. Tranfield D, Denyer D, Smart P. Towards a Methodology for Developing Evidence-Informed Management Knowledge by Means of Systematic Review. British Journal of Management 2003 Sep 1 [cited 2023 Jul 13];14(3):207–22. doi: 10.1111/1467-8551.00375

22. Snyder H. Literature review as a research methodology: An overview and guidelines. J Bus Res. 2019 Nov 1;104:333–9.

23. Page MJ, McKenzie JE, Bossuyt PM, Boutron I, Hoffmann TC, Mulrow CD, et al. The PRISMA 2020 statement: an updated guideline for reporting systematic reviews. Syst Rev 2021 Dec 1 [cited 2023 Jul 13];10(1):1–11. doi: 10.1186/s13643-021-01626-4

24. Umesh UN, Huynh MQ, Jessup L. Creating successful entrepreneurial ventures in IT. Commun ACM. 2005 Jun;48(6):82–7.

25. Long HA, French DP, Brooks JM. Optimising the value of the critical appraisal skills programme (CASP) tool for quality appraisal in qualitative evidence synthesis. 2020;1(1):31–42. doi: 10.1177/2632084320947559

26. McCliskey, Annie, & O’Connor D. Implementing evidence: Closing research–practice gaps-ClinicalKey. Hoffmann, Tammy, Bennett, Sally, & Del MArC, editor. Evidence-Based Practice Across Health Professions 2017;384–408. ISBN ebook: 9780729586085

27. Hannes K, Macaitis K. A move to more systematic and transparent approaches in qualitative evidence synthesis: Update on a review of published papers. Qualitative Research 2012;12(4):402–42. doi: 10.1177/1468794111432992

28. Gompers P, Kovner A, Lerner J, Scharfstein D. Performance persistence in entrepreneurship. J financ econ 2010 Apr;96(1):18–32. doi: 10.1016/j.jfineco.2009.11.001

